# Chemotherapy use and outcomes in patients with stage III or IV small-cell lung cancer in relation to age: An analysis of the English Systemic Anti-Cancer Treatment (SACT) dataset

**DOI:** 10.1101/2022.07.06.22277250

**Authors:** S. Pilleron, EJA. Morris, D. Dodwell, K.N. Franks

## Abstract

**Introduction:** Survival from small cell lung cancer (SCLC) is poor and there has been little progress in treatment. There is little evidence on outcomes in patients aged 75+. We described patterns of chemotherapy use and outcomes using the national Systemic Anti-Cancer Treatment dataset in patients with stage III or IV SCLC in relation to age.

**Method:** We included 7,966 SCLC (67.6% stage IV) diagnosed between 2014-17 in England, treated with chemotherapy and followed up through 2017. Patterns of chemotherapy use, 30- and 90-mortality rates, 6-,12-month and median overall survival (OS) from the start of the first chemotherapy cycle were compared between those below and above the age of 75. OS was estimated using Kaplan Meier estimator and modeled using a flexible hazard regression model.

**Results:** Older patients were 6-7 times less likely to receive curative treatment than younger patients regardless of stage. There were more frequent adjustments of treatment and dose reduction (stage III) in older than younger patients but no age-related differences in reduction of doses (stage IV), treatment delayed or stopped earlier than planned. Although 30-day mortality rates were similar across age groups in stage III SCLC (∼4%), older patients had higher early mortality and poorer OS than younger peers. In both stages, 6 and 12-month OS by age decreased around the age of 70-75 and were worse in patients with performance status scores ≥2.

**Conclusion:** This study offers a snapshot of chemotherapy use and outcomes in advanced SCLC, notably in older patients, in the pre-immunotherapy era.

## Introduction

Lung cancer affects more than 45,000 people and causes more than 35,000 deaths in the United Kingdom (UK) each year. ^1^ Whilst standardised incidence rates are decreasing, the absolute numbers of new lung cancer cases and deaths attributed to lung cancer are expected to increase due to an ageing population. ^2^

Small cell lung cancer (SCLC) represent about 10-15% of all lung cancer, ^3^ and the majority of patients are diagnosed at an advanced stage. ^4^ Survival from SCLC is poor, and the UK has reported lower survival rates than other high-income countries with similar healthcare systems. ^4^

While chemotherapy is the accepted mainstay of treatment for SCLC, there is no direct randomized evidence of its superiority over supportive care. ^5^ There has been little progress with new cytotoxic chemotherapy agents or combinations over many years though recent trials have shown modest survival benefits with the addition of immunotherapy to platinum based chemotherapy in extensive disease. ^6,7^

Adults aged 75 years or older are not commonly included in randomized clinical trials because of comorbidities, poor performance status, eligibility criteria, or patient preference. ^8^ Older patients are also less likely to receive active treatment and may be at higher risk of toxicity and early mortality. ^9^ Little is known about the outcomes for older people who do receive treatment. ^9^

The UK NICE guidelines recommend the use of cytotoxic chemotherapy in incurable advanced/metastatic disease for patients with good performance status. There are no specific recommendations with regard to age. ^10^ To our knowledge, there is no description of current practice of chemotherapy use in SCLC in England in relation to age.

The Systemic Anti-Cancer Treatment (SACT) dataset is a national population-based dataset that collects data on SACT treatment (including chemotherapy, immunotherapy, targeted therapy, or combination therapy) in all patients with cancer. ^11^ This study sought to use this resource to describe patterns of chemotherapy use, and associated outcomes in patients diagnosed with stage III or IV SCLC in relation to age prior to the NICE approval of immunotherapy given with platinum-based chemotherapy in extensive stage SCLC.

## Materials and Methods

In this retrospective observational population-based study, we included patients diagnosed with stage III or stage IV SCLC (International Classification of Diseases for Oncology 2^nd^ edition (ICD-O-2): 8002, 8041, 8042, 8043,8044, 8045) between 1^st^ January 2014 and 31^st^ December 2017 aged 18+ who received chemotherapy. We restricted our analyses to patients who were recorded as receiving chemotherapy alone for lung cancer (supplemental Table 1) in the SACT dataset as this was the treatment recommended by national guidelines for extensive SCLC. ^10^ Patient (age, sex, ethnicity, socio-economic deprivation level [measured using quintiles of the income domain of the Index of Multiple Deprivation (IMD)] ^12^ and tumour (morphology, stage at diagnosis) characteristics as well as vital status and date of death were retrieved from the National Cancer Registration and Analysis Service (NCRAS) data linked to SACT. We excluded cases diagnosed based on death certificates only, or a second lung cancer diagnosed within the six first months after the first diagnosis. We further excluded records for patients who took part in a trial, or were recorded as receiving “not chemo”, “not matched’ or “missing treatment group”, duplicate records, records with treatment dates before 1^st^ January 2014 or after 31^st^ December 2018, or when treatment date was reported after date of death. Vital status was updated on 31 December 2018.

We retrieved information recorded about chemotherapy regimens and associated outcomes, and chemotherapy cycles. Regimen data included start date, treatment intent, treatment given, whether the regimen was adapted based on comorbidities (choice of regimen, dose modification or change in treatment interval). Adjuvant and neoadjuvant treatments were categorised into curative treatment. A cancer clinician (KNF) checked for inconsistencies between chemotherapy regimen and treatment intent. In case of inconsistencies, intent was recoded based on review of the regimes used in the context of SCLC management (supplemental Table 2). If a chemotherapy regimen may be used in palliative or curative intent (i.e. Cisplatin, EC, EP), we kept the original coding. If the original coding was “disease modification”, we kept as is.

Outcome data included ^11^:

- Dose reduction = Identifies if a regimen was recorded as being modified by reducing the dose of any anti-cancer drug administered at any point in the regimen.
- Time delay = Identifies if a regimen was recorded as being modified by extending the time between administration dates at any point in the regimen.
- Regimen stopped early = Identifies if a regimen was recorded as being modified by reducing the administration days below the number planned.

Data on cycle comprised start date, and performance status at start of cycle. We recoded performance status into 0,1, 2+ and missing and we used the performance status recorded at the start of the first chemotherapy cycle in the analysis.

We calculated the number of regimens and cycles each patient received. We also created a variable to indicate whether patients who received a first regimen with curative intent received chemotherapy with recorded palliative intent subsequently.

### Statistical analysis

All analyses were stratified by stage at diagnosis. Descriptive statistics were applied as appropriate: percentages for categorical variables and medians with its interquartile ranges for continuous variables.

We estimated the mortality rates at 30- and 90-days from the start of the first cycle of chemotherapy.

We used the Kaplan-Meier estimator to estimate the 6-month and 1-year overall survival (OS) and median survival time with its 95% confidence interval from the start of the 1^st^ cycle of chemotherapy in younger and older patients by stage and performance status. The end of follow-up was 31^st^ December 2018.

To further describe overall survival by age, we derived, for each stage at diagnosis, overall survival from the estimation of individual mortality hazard using flexible hazard regression models. We modelled the hazard function as the exponential of B-spline of degree 3 with a knot located at the median of the distribution of survival times in patients who died. We included age as a B-spline of degree 3 with a knot at the median of the distribution of age in the whole sample. We first ran models including treatment intent of the first regimen as a factor, and then models including performance status as a factor.

We performed statistical analyses using R statistical software (version 3.4.0; R Development Core Team, 2017). We used the ‘mexhaz’ package to model hazard and estimate survival. ^13^

### Ethics

Approval for the study was granted by Public Health England Office for Data Release (Reference ODR/1920/080) and North of Scotland Research Ethics Service gave ethical approval for this work (REC/19/NS/0057). Informed consent from individual participants was not required.

## Results

Out of 28,698 lung cancer cases diagnosed in 2014-2017 recorded as having received SACT, we retained 7,966 SCLC (2,580 stage III cancer and 5,386 stage IV) cases for the present study. The flow chart is presented in Figure 1.

**Figure 1.**
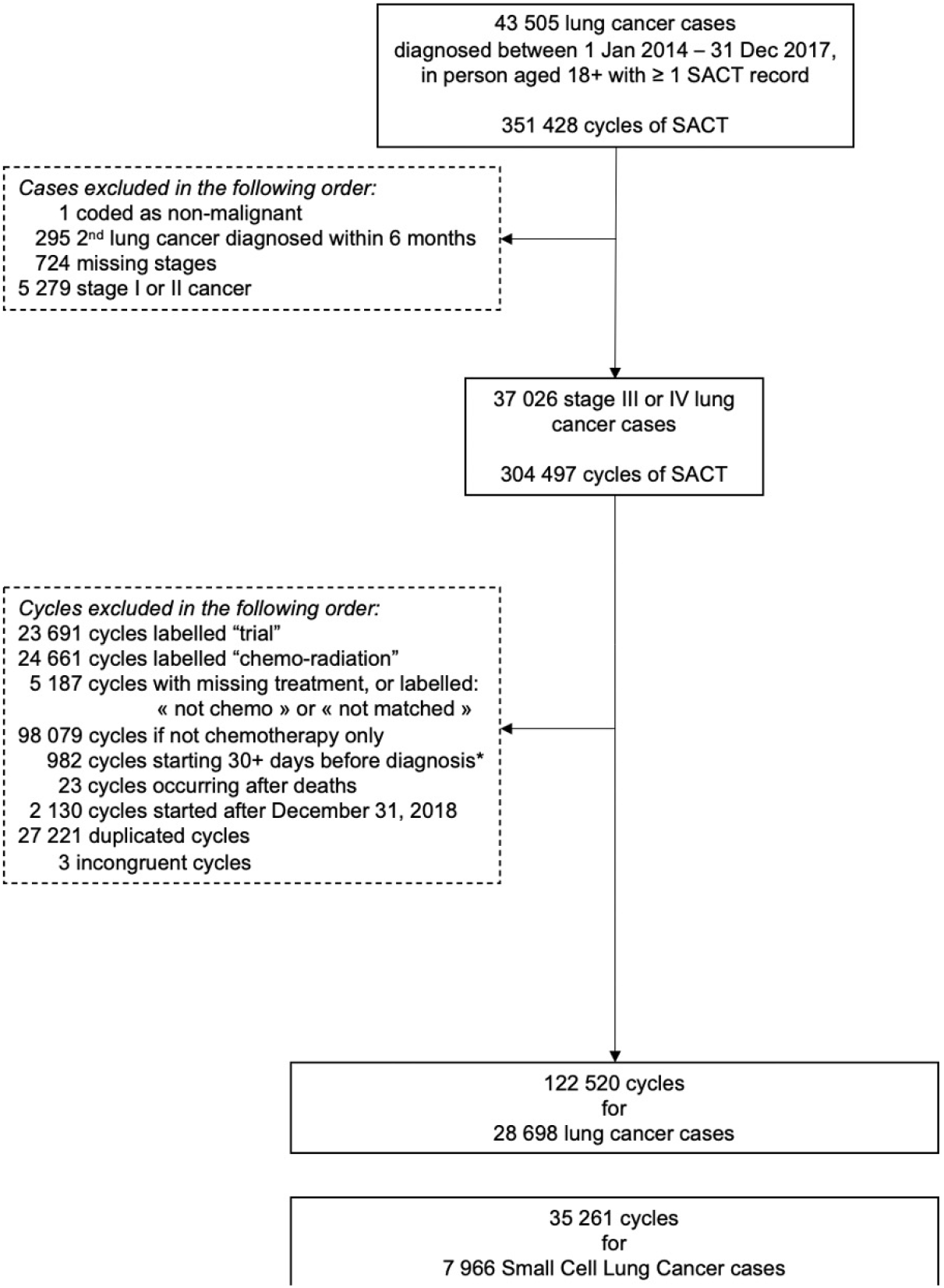
Flow chart Note. ^*^ We allowed chemotherapy to start within 30 days prior to cancer diagnosis to consider possible differences between the date of cancer clinically diagnosed and the date of cancer diagnosis recorded in the registry.

The distribution of age at diagnosis across stages was similar for both stages III and IV with a median age of 68 years (Figure 2). Females comprised 55.1% and 47.8% of patients with stage III and stage IV, respectively.

**Figure 2.**
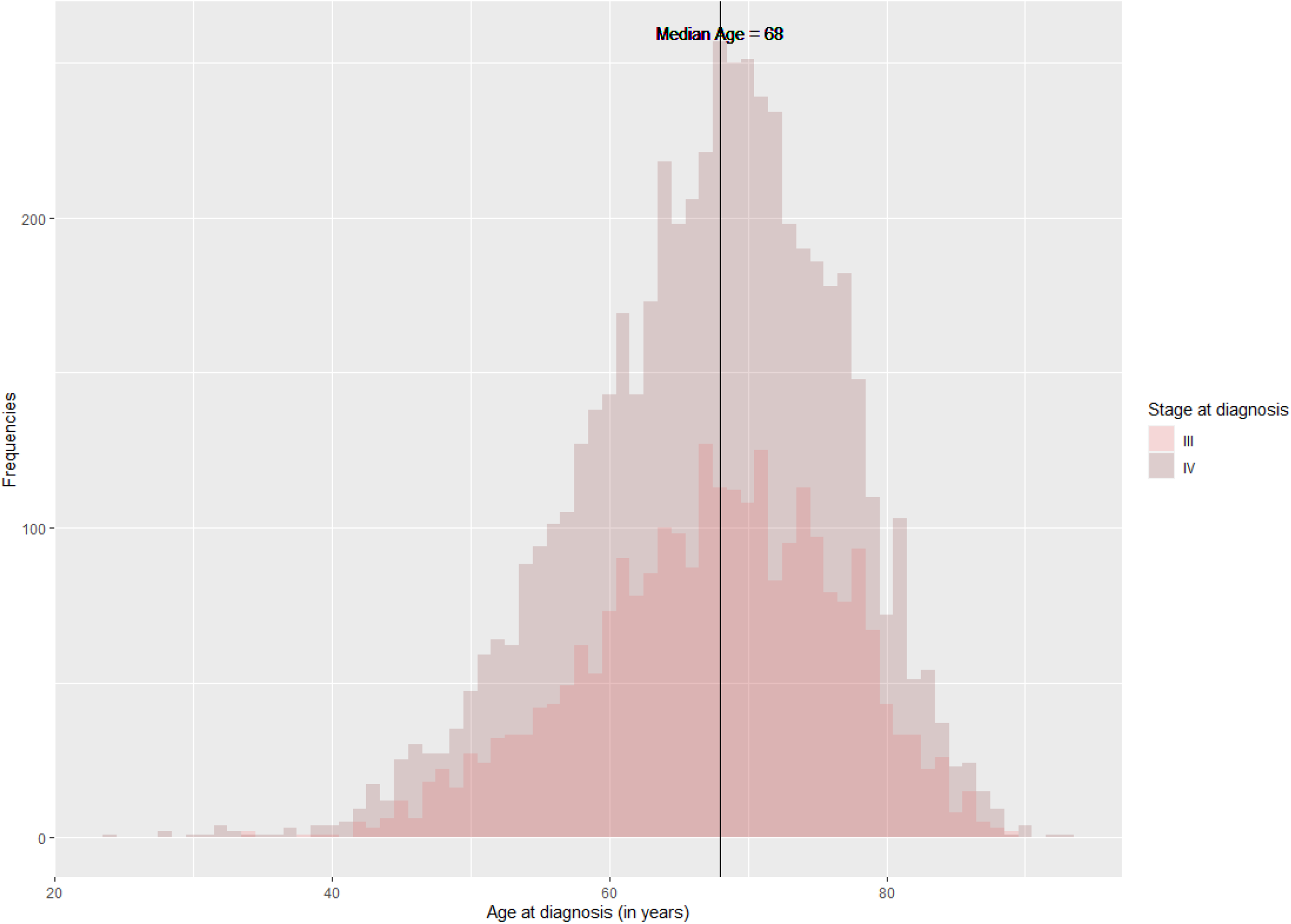
Distribution of age at diagnosis of patients diagnosed with stage III or IV SCLC in 2014-2017 who received chemotherapy

Table 1 describes patients’ characteristics based on stage in patients below and above 75 years old. Overall, 23% of patients with stage III SCLC and 22% with stage IV disease were aged ≥75, and ∼7% were aged ≥80 regardless of stage. Older patients had lower performance status scores than younger patients in both stages. While most patients with SCLC received palliative treatment, older adults were 6-7 times less likely to receive curative treatment than younger patients regardless of stage (3.3% vs 20.1% in stage III and 0.8% vs 7.3% in stage IV). This is also evident when looking at the age distribution by stage and treatment intent (supplemental Figure 1). Patients aged over 75 were much less represented in patients receiving curative treatment than in those receiving palliative treatment. Regardless of age group or stage, patients with SCLC received a median number of 4 cycles.

**Table 1.** Characteristics of patients diagnosed with stage III or IV small cell lung cancer in England in 2014-2017 and who received chemotherapy by age group

|  | Stage III |  | Stage IV |  |
| --- | --- | --- | --- | --- |
|  | <75 years old | 75+ years old | <75 years old | 75+ years old |
| n | 1978 | 602 | 4187 | 1199 |
| Median age at diagnosis (years [IQR]) | 65.0<br>[60.0, 70.0] | 78.0<br>[76.0, 80.0] | 65.0<br>[59.0, 70.0] | 78.0<br>[76.0, 81.0] |
| Females - n (%) | 1121 (56.7) | 301 (50.0) | 2075 (49.6) | 501 (41.8) |
| Ethnicity - n (%) |  |  |  |  |
| White | 1890 (95.6) | 567 (94.2) | 3936 (94.0) | 1144 (95.4) |
| Non-White | 55 (2.8) | 16 (2.7) | 120 (2.9) | 30 (2.5) |
| Unknown/Unstated | 33 (1.7) | 19 (3.2) | 131 (3.1) | 25 (2.1) |
| Performance status<br>at start of the 1st cycle - n (%) |  |  |  |  |
| 0 | 465 (23.5) | 91 (15.1) | 633 (15.1) | 123 (10.3) |
| 1 | 726 (36.7) | 228 (37.9) | 1436 (34.3) | 371 (30.9) |
| 2+ | 355 (17.9) | 168 (27.9) | 1212 (28.9) | 461 (38.4) |
| Missing | 432 (21.8) | 115 (19.1) | 906 (21.6) | 244 (20.4) |
| Treatment intent of the 1<sup>st</sup> regimen<br>- n (%) |  |  |  |  |
| Curative | 392 (19.8) | 20 (3.3) | 302 (7.2) | 10 (0.8) |
| Palliative | 1560 (78.9) | 573 (95.2) | 3851 (92.0) | 1182 (98.6) |
| Disease modification | 26 (1.3) | 9 (1.5) | 34 (0.8) | 7 (0.6) |
| Change from curative to palliative intent among those who received curative treatment for their 1<sup>st</sup> regimen - n (%) | 213 (54.3) | 18 (90.0) | 177 (58.6) | 7 (70.0) |
| Median number of cycles recorded [IQR] | 4.0 [3.0, 6.0] | 4.0 [3.0, 5.0] | 4.0 [2.0, 6.0] | 4.0 [1.0, 5.0] |

IQR: Interquartile range

### Change of treatment plans

The first regimen of chemotherapy was adapted as a result of the presence of comorbidities more often in older than younger patients regardless of stage (∼26% vs 20% - Table 2). Chemotherapy doses were reduced more often in older compared to younger patients with stage III SCLC (35% vs 29%). However, the percentages of cases that underwent a reduction of doses in stage IV, had delayed treatment or stopped earlier than planned were similar in both age groups in both stages.

**Table 2.** Changes in treatment plans by age group and stage at diagnosis

|  | Stage III |  | Stage IV |  |
| --- | --- | --- | --- | --- |
|  | <75 | ≥75 | <75 | ≥75 |
| ***First regimen recorded*** |  |  |  |  |
| Adjusted for comorbidity - n (% of non-missing) | 293 (21.1) | 119 (26.6) | 648 (20.3) | 247 (26.3) |
| Missing – n (% total) | 588 (29.7) | 155 (25.7) | 991 (23.7) | 260 (21.7) |
| Dose reduction - n (% of non-missing) |  |  |  |  |
| Yes | 442 (28.6) | 169 (35.4) | 1083 (31.9) | 325 (33.4) |
| Missing – n (% total) | 431 (21.8) | 124 (20.6) | 790 (18.9) | 227 (18.9) |
| Time delay - n (% of non-missing) |  |  |  |  |
| Yes | 380 (35.0) | 123 (35.9) | 799 (32.7) | 243 (34.2) |
| Missing – n (% total) | 893 (45.1) | 259 (43.0) | 1742 (41.6) | 488 (40.7) |
| Stopped earlier - n (% of non-missing) |  |  |  |  |
| Yes | 218 (15.4) | 74 (16.4) | 491 (15.4) | 137 (14.7) |
| Missing – n (% total) | 561 (28.4) | 152 (25.2) | 993 (23.7) | 269 (22.4) |
| ***In any subsequent regimens recorded*** |  |  |  |  |
| # patients with >1 regimen recorded | 1166 | 306 | 2167 | 477 |
| Adjustment for comorbidity - n (% of non-missing) | 173 (21.4) | 66 (30.4) | 349 (21.6) | 102 (28.1) |
| Missing – n (% total) | 357 (30.6) | 89 (29.1) | 554 (25.6) | 114 (23.9) |
| Dose reduction (% of non-missing) | 350 (37.9) | 98 (41.4) | 715 (40.9) | 172 (45.3) |
| Missing – n (% total) | 242 (20.8) | 69 (22.5) | 417 (19.2) | 97 (20.3) |
| Time delay - (% of non-missing) | 296 (42.3) | 77 (41.6) | 552 (41.3) | 153 (48.6) |
| Missing – n (% total) | 467 (40.1) | 121 (39.5) | 831 (38.3) | 162 (34.0) |
| Stopped earlier - (% of non-missing) | 175 (20.4) | 43 (18.8) | 346 (21.0) | 62 (17.3) |
| Missing – n (% total) | 309 (26.5) | 77 (25.2) | 516 (23.8) | 118 (24.7) |

Regarding subsequent regimens, adjustments of treatment because of comorbidities were also observed more often in older patients than in younger ones (∼28-30% for older patients for both stages vs ∼21% for younger patients). Similar findings were seen for dose reductions: 41% in older patients with stage III SCLC vs 38% in younger patients and 45% vs 41% respectively in patients with stage IV disease. The delay between cycle was modified in >40% of patients regardless of age group and stage. Finally, percentages of patients who had their regimen stopped earlier than planned were similar in both age groups regardless of stage (∼20%).

### Early mortality rates and survival

Table 3 presents early mortality and survival outcomes in older and younger patients by stage. While there is no difference in 30-day mortality rates across age groups in patients with stage III SCLC, older patients had higher mortality rates with the biggest difference observed in older patients with stage IV (28% vs 20%). They also had poorer 6 and 12-month survival than younger patients with differences of 7 to 11.5 percentage points, and a 2-month shorter median survival regardless of stage.

**Table 3.**
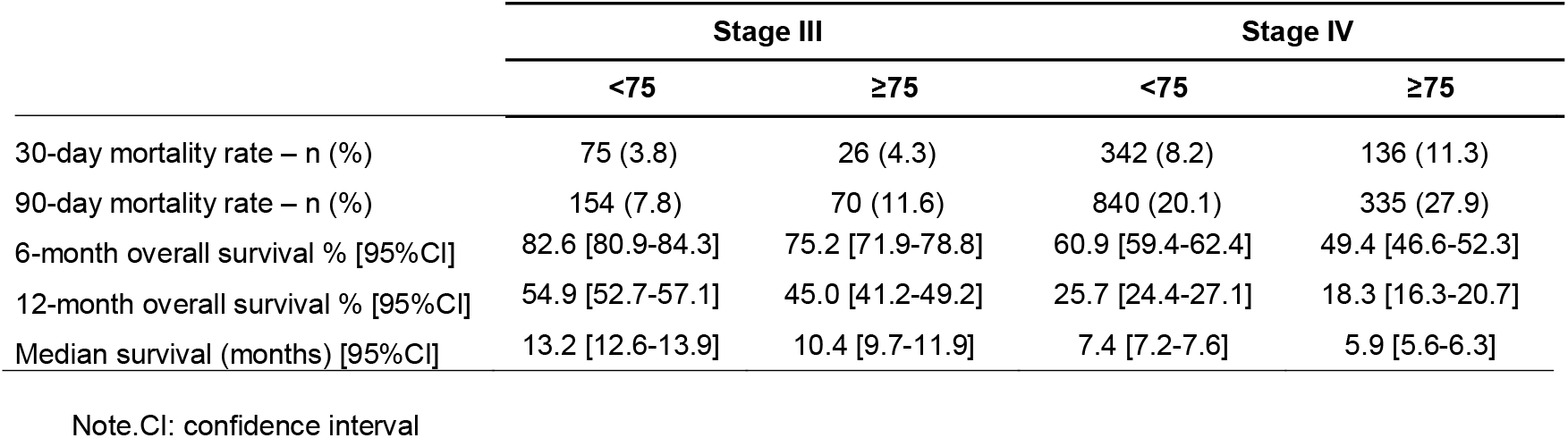
30-and 90-day mortality rates, 6-and 12-month and median overall survival from the start of the 1^st^ cycle of chemotherapy by stage in small cell lung cancer cases aged below and above 75 treated with chemotherapy

|  | Stage III |  | Stage IV |  |
| --- | --- | --- | --- | --- |
| | <75 | $\geq 75$ | <75 | $\geq 75$ |
| 30-day mortality rate – n (%) | 75 (3.8) | 26 (4.3) | 342 (8.2) | 136 (11.3) |
| 90-day mortality rate – n (%) | 154 (7.8) | 70 (11.6) | 840 (20.1) | 335 (27.9) |
| 6-month overall survival % [95%CI] | 82.6 [80.9-84.3] | 75.2 [71.9-78.8] | 60.9 [59.4-62.4] | 49.4 [46.6-52.3] |
| 12-month overall survival % [95%CI] | 54.9 [52.7-57.1] | 45.0 [41.2-49.2] | 25.7 [24.4-27.1] | 18.3 [16.3-20.7] |
| Median survival (months) [95%CI] | 13.2 [12.6-13.9] | 10.4 [9.7-11.9] | 7.4 [7.2-7.6] | 5.9 [5.6-6.3] |

Note.CI: confidence interval

When stratified by performance status and stage (supplemental Table 3), early mortality and overall survival were poorest in patients with a performance status score ≥2, while similar in those with performance status score of 0 and 1 in both age groups and stages. Differences in mortality rates and survival across age groups are smaller in patients with stage III SCLC than in those with stage IV disease.

To better describe how age affects survival we looked at the age pattern of overall survival using age as a continuous variable over the initial year from the start of the first chemotherapy (Figure 3). Regardless of stage or treatment intent, overall survival decreased over time as age increased (Figure 3 - A). Survival at 6 and 12-month by age (Figure 3 – B) decreased around the age of 70-75 in both stages. Finally, Figure 3C shows that overall survival at month 6 did not differ much between a performance status score of 0 or 1 but was significantly worse for a score of ≥2 in all ages. 6-month overall survival in patients with missing performance status was in-between patients with score of 1 and those with a score ≥2.

**Figure 3.**
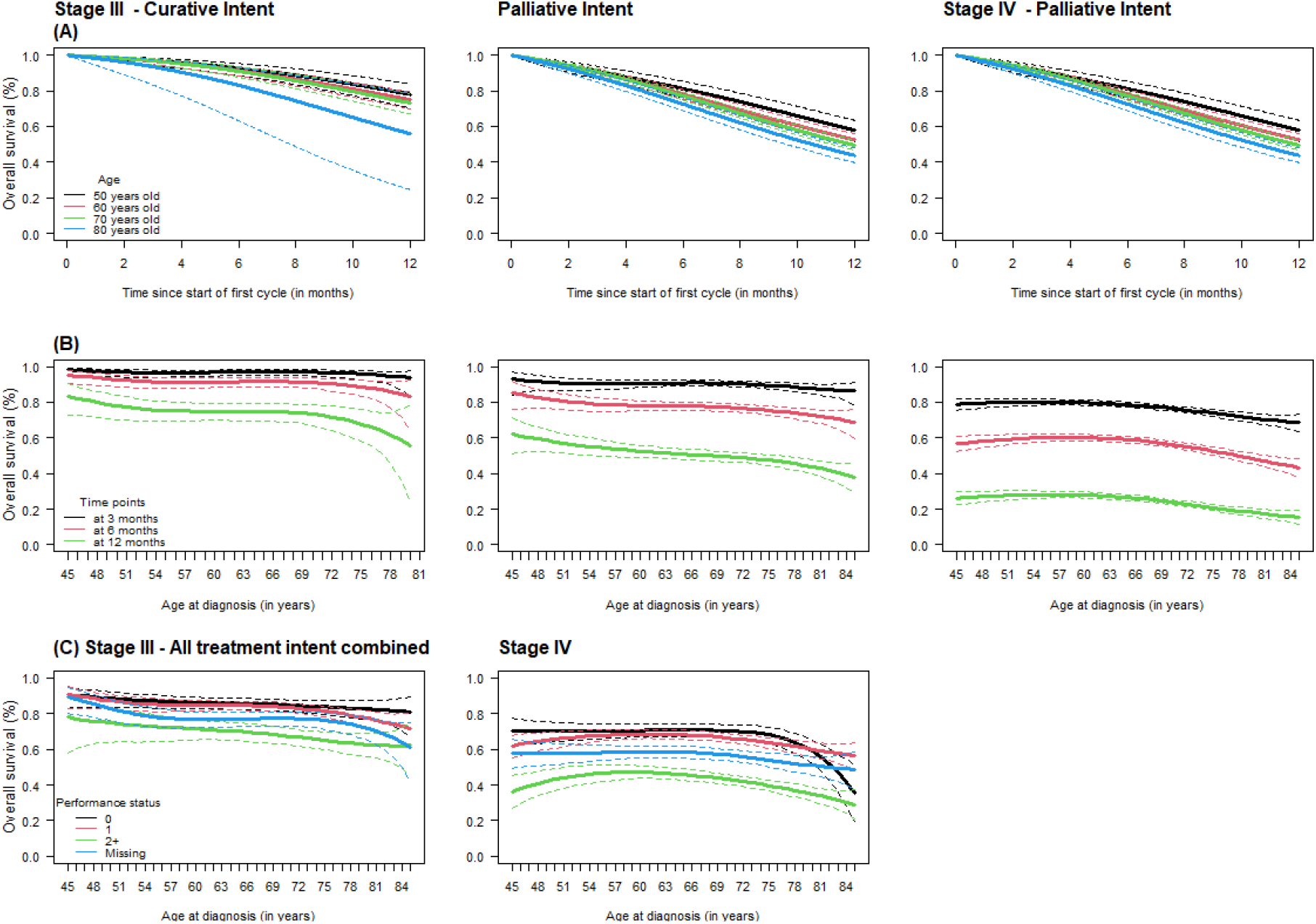
Overall survival (OS) over the first year from the start of the first cycle of chemotherapy at ages 50, 60, 70, and 80; (B) 6, and 12-month OS by age at diagnosis and (C) 6-month OS by age and performance status in patients diagnosed with stage III and IV small cell lung cancer years old diagnosed in 2014-2017 and treated with chemotherapy

## Discussion

This observational population-based study is the first to describe the pattern of chemotherapy use and associated outcomes in relation to age in patients diagnosed with SCLC and treated with chemotherapy in England. Despite a similar pattern of use of chemotherapy across age groups, the 30-day mortality rate in stage IV SCLC and overall survival were poorer as age increased, notably from 70-75 years old. Even though the study covered the pre-immunotherapy period, this study is relevant to contemporary care as patients need to be fit enough to receive immunotherapy and chemotherapy alone is still a commonly used treatment option in patients with poor performance status (PS ≥2) or those with a contraindication to immunotherapy. In addition, the benefit of adding immunotherapy is relatively small with a median overall survival improvement of 2 months with Atezolizumab ^14,15^ and <3 months with Durvalumab. ^16,17^

Outside England, few hospital-based studies described treatment patterns in patients diagnosed with small cell lung cancer. ^18–21^ Among them, only a Chinese study including patients diagnosed with SCLC from 1996 to 2005 addressed differences in outcomes between patients receiving cyclophosphamide, adriamycin and vincristine vs. etoposide and cisplatin, aged below and above 70 years. ^20^ Two population-based studies conducted in Canada and the US described chemotherapy use in patients diagnosed with SCLC but did not stratify their results by age. ^22,23^ A recent review of real-world evidence of treatment patterns and outcomes reported that these had remained poor over the preceding 20 years but they did not present their results by age. ^24^

Our results should be interpreted considering contemporary practice. The calendar period of the study pre-dated the routine use of immunotherapy. During the study period, national guidelines recommended cytotoxic chemotherapy in incurable advanced/metastatic disease for patients assessed as fit enough, with no specific recommendations for older patients. ^10^ Because of the underrepresentation of adults aged 75 years or older in clinical trials, chemotherapy is offered without strong evidence about the likely benefits and risks in this population. In our study, older patients have comparable outcomes to younger patients, suggesting that these patients were selected appropriately for active treatment.

The present study is nationally inclusive, based on records of all patients diagnosed with SCLC in the English National Health Service (NHS) who were recorded to have had chemotherapy alone, representing ∼60% of all patients diagnosed with SCLC during 2014-2017 period (based on an estimate of ∼3800 SCLC per year ^4^). Follow-up was adequate to address outcomes in SCLC and data completeness for stage and treatment intent was high. However, our study has limitations. Data may still be incorrectly coded and source verification was not possible. Data were unavailable or insufficiently complete on treatment response, disease progression, and co-morbidities so these outcomes could not be assessed.

## Conclusion

Using national population-based data on chemotherapy in England, we describe for the first time the chemotherapy patterns and associated outcomes in patients diagnosed with SCLC between 2014 and 2017 in relation to age. Despite similar chemotherapy treatments, older adults have poorer survival outcomes and are at high risk of early mortality compared to younger patients. Further information is required to optimise the treatment for older patients, particularly in the era of immunotherapy and combination treatment, but the outcomes we report can be used in appropriate circumstances to share information and support shared decision-making.

## Data Availability

De-personalised study data may be made available on request to accredited researchers who submit a proposal that is approved by the PHE Office for Data Release.

## Acknowledgements

We thank Dr. Zhe Wang for his statistical help and James Thomas NHS-Digital for help with data acquisition, interpretation, and processing.

## List of supplementary material

### Tables

Supplemental Table 1 - Regimen coding

Supplemental Table 2 – Regimen associated to their treatment intent

Supplemental Table 3 - 30-and 90-day mortality rates and 6-and 12-month and median overall survival from the start of the 1^st^ cycle of chemotherapy by stage and performance status in small cell lung cancer cases aged below and above 75 and treated with chemotherapy

### Figure

Supplemental Figure 1 – Age distribution by stage and treatment intent

