## Supplemental Tables and Figure for "Chemotherapy use and outcomes in patients with stage III or IV small-cell lung cancer in relation to age: An analysis of the English Systemic Anti-Cancer Treatment (SACT) dataset"

---

**List of supplementary material**

Tables

Supplemental Table 1 - Regimen coding

Supplemental Table 2 – Regimen associated to their treatment intent

Supplemental Table 3 - 30-and 90-day mortality rates and 6-and 12-month and median overall survival from the start of the 1st cycle of chemotherapy by stage and performance status in small cell lung cancer cases aged below and above 75 and treated with chemotherapy

Figure

Supplemental Figure 1 – Age distribution by stage and treatment intent

Supplemental Table 1 - Regimen coding

| Regimen | Category | Frequencies |
|---|---|---|
| CARBOPLATIN | Chemotherapy alone | 2375 |
| CARBOPLATIN + DOCETAXEL | Chemotherapy alone | 12 |
| CARBOPLATIN + ETOPOSIDE | Chemotherapy alone | 39339 |
| CARBOPLATIN + GEMCITABINE + PACLITAXEL | Chemotherapy alone | 5 |
| CARBOPLATIN + IRINOTECAN | Chemotherapy alone | 16 |
| CARBOPLATIN + NAB-PACLITAXEL | Chemotherapy alone | 12 |
| CARBOPLATIN + PACLITAXEL | Chemotherapy alone | 1878 |
| CARBOPLATIN + PEMETREXED | Chemotherapy alone | 20721 |
| CARBOPLATIN + TOPOTECAN | Chemotherapy alone | 1 |
| CARBOPLATIN + VINORELBINE | Chemotherapy alone | 10590 |
| CYCLOPHOSPHAMIDE + DOXORUBICIN + VINCRISTINE | Chemotherapy alone | 2080 |
| CISPLATIN | Chemotherapy alone | 132 |
| CISPLATIN + DOCETAXEL | Chemotherapy alone | 47 |
| CISPLATIN + DOXORUBICIN | Chemotherapy alone | 4 |
| CISPLATIN + ETOPOSIDE | Chemotherapy alone | 3897 |
| CISPLATIN + GEMCITABINE | Chemotherapy alone | 1833 |
| CISPLATIN + PACLITAXEL | Chemotherapy alone | 13 |
| CISPLATIN + PEMETREXED | Chemotherapy alone | 15513 |
| CISPLATIN + VINCRISTINE | Chemotherapy alone | 4 |
| CISPLATIN + VINORELBINE | Chemotherapy alone | 7064 |
| DOCETAXEL | Chemotherapy alone | 3909 |
| EC (ETOPOSIDE + PLATINUM (CISPLATINE or CARBOPLATIN)) | Chemotherapy alone | 5 |
| EP (ETOPOSIDE + PLATINUM (CISPLATINE or CARBOPLATIN)) | Chemotherapy alone | 35 |
| ETOPOSIDE | Chemotherapy alone | 83 |
| EVEROLIMUS | Chemotherapy alone | 151 |
| GEMCARBO | Chemotherapy alone | 22079 |
| GEMCITABINE | Chemotherapy alone | 467 |
| PACLITAXEL | Chemotherapy alone | 361 |
| PACLITAXEL + PEMETREXED | Chemotherapy alone | 7 |
| PEMETREXED | Chemotherapy alone | 17931 |
| TOPOTECAN | Chemotherapy alone | 1345 |
| VINORELBINE | Chemotherapy alone | 970 |
| CARBOPLATIN + PEMBROLIZUMAB + PEMETREXED | Chemotherapy + Immunotherapy | 3 |
| ATEZOLIZUMAB + BEVACIZUMAB + CARBOPLATIN + PACLITAXEL | Chemotherapy + Immunotherapy + Targeted Therapy | 31 |
| ATEZOLIZUMAB | Immunotherapy alone | 3714 |
| DURVALUMAB | Immunotherapy alone | 289 |
| IPILIMUMAB | Immunotherapy alone | 2 |
| IPILIMUMAB + NIVOLUMAB | Immunotherapy alone | 7 |
| NIVOLUMAB | Immunotherapy alone | 943 |
| PEMBROLIZUMAB | Immunotherapy alone | 44228 |
| DENOSUMAB | Not chemotherapy - bone | 8581 |
| PAMIDRONATE | Not chemotherapy - bone | 46 |
| ZOLEDRONIC ACID | Not chemotherapy - bone | 2850 |

| | | |
|---|---|---|
| ABIRATERONE | Not Lung | 7 |
| ACE | Not Lung | 61 |
| AFLIBERCEPT + FU + IRINOTECAN | Not Lung | 2 |
| BEP | Not Lung | 2 |
| BEVACIZUMAB + IRINOTECAN + MDG | Not Lung | 3 |
| BORTEZOMIB | Not Lung | 4 |
| CABOZANTINIB | Not Lung | 5 |
| CAPECITABINE | Not Lung | 50 |
| CAPECITABINE + CARBOPLATIN | Not Lung | 4 |
| CAPECITABINE + CARBOPLATIN + EPIRUBICIN | Not Lung | 6 |
| CAPECITABINE + CISPLATIN | Not Lung | 8 |
| CAPECITABINE + DOCETAXEL | Not Lung | 1 |
| CAPECITABINE + EPIRUBICIN + OXALIPLATIN | Not Lung | 15 |
| CAPECITABINE + GEMCITABINE | Not Lung | 43 |
| CAPECITABINE + OXALIPLATIN | Not Lung | 15 |
| CAPECITABINE + STREPTOZOCIN | Not Lung | 26 |
| CAPECITABINE + TEMOZOLOMIDE | Not Lung | 91 |
| CAPECITABINE + VINORELBINE | Not Lung | 3 |
| CARBO + FLUOROURACIL | Not Lung | 5 |
| CARBOPLATIN + CETUXIMAB + FLUOROURACIL | Not Lung | 4 |
| CARBOPLATIN + CETUXIMAB + FU | Not Lung | 3 |
| CARBOPLATIN + EPIRUBICIN | Not Lung | 12 |
| CETUXIMAB | Not Lung | 8 |
| CETUXIMAB + CISPLATIN + FU | Not Lung | 2 |
| CHOP | Not Lung | 11 |
| CHOP R | Not Lung | 12 |
| CISPLATIN + FLUORO + STREPTOZOCIN | Not Lung | 3 |
| CISPLATIN + FLUOROURACIL | Not Lung | 5 |
| CTD | Not Lung | 5 |
| CVD | Not Lung | 5 |
| CVP | Not Lung | 1 |
| CVP R | Not Lung | 6 |
| CVP R + GEMCITABINE | Not Lung | 6 |
| CYCLO + DOXORUBICIN + VINCRISTINE | Not Lung | 209 |
| CYCLOPHOSPHAMIDE | Not Lung | 1 |
| CYCLOPHOSPHAMIDE + DOXORUBICIN + ETOPOSIDE | Not Lung | 5 |
| CYCLOPHOSPHAMIDE + ETOPOSIDE | Not Lung | 26 |
| CYTARABINE | Not Lung | 3 |
| DOCETAXEL + PERTUZUMAB + TRASTUZUMAB | Not Lung | 7 |
| DOCETAXEL + TRASTUZUMAB | Not Lung | 2 |
| ECF | Not Lung | 16 |
| ECX | Not Lung | 51 |
| EMA | Not Lung | 1 |
| ENZALUTAMIDE | Not Lung | 3 |
| EOX | Not Lung | 10 |
| EPIRUBICIN | Not Lung | 2 |
| ETOPOSIDE + LOMUSTINE + VINCRISTINE | Not Lung | 17 |

| | | |
|---|---|---|
| FCARBOST | Not Lung | 15 |
| FCIST | Not Lung | 1 |
| FEC | Not Lung | 7 |
| FLUOROURACIL | Not Lung | 1 |
| FLUOROURACIL + STREPTOZOCIN | Not Lung | 1 |
| HCX | Not Lung | 5 |
| HORMONES | Not Lung | 115 |
| HYDROXYCARBAMIDE | Not Lung | 3 |
| IMATINIB | Not Lung | 13 |
| IPM | Not Lung | 60 |
| IRINOTECAN | Not Lung | 1 |
| IRINOTECAN + MDG | Not Lung | 29 |
| LANREOTIDE | Not Lung | 198 |
| MCX | Not Lung | 1 |
| METHOTREXATE HIGH DOSE | Not Lung | 3 |
| MITOMYCIN INTRAVESICULAR | Not Lung | 4 |
| MVP | Not Lung | 1 |
| OCTREOTIDE | Not Lung | 93 |
| OXALIPLATIN + MDG | Not Lung | 57 |
| OXALIPLATIN + MDG + PANITUMUMAB | Not Lung | 2 |
| OXALIPLATIN + RALTITREXED | Not Lung | 8 |
| PACLITAXEL + TRASTUZUMAB | Not Lung | 4 |
| PAZOPANIB | Not Lung | 2 |
| PERTUZUMAB + TRASTUZUMAB | Not Lung | 24 |
| RALTITREXED | Not Lung | 1 |
| RITUXIMAB | Not Lung | 14 |
| RUXOLITINIB | Not Lung | 4 |
| STREPTOZOCIN + MDG | Not Lung | 8 |
| TEMOZOLOMIDE | Not Lung | 10 |
| TRASTUZUMAB | Not Lung | 8 |
| VIP | Not Lung | 2 |
| ATEZOLIZUMAB + BEVACIZUMAB | Targeted Therapy + immunotherapy | 3 |
| DOCETAXEL + NINTEDANIB | Targeted Therapy + immunotherapy | 4159 |
| BEVACIZUMAB + CARBOPLATIN + PACLITAXEL | Targeted Therapy + chemotherapy | 2 |
| CARBO + GEFITINIB + PEMETREXED | Targeted Therapy + chemotherapy | 3 |
| AFATINIB | Targeted Therapy alone | 9462 |
| ALECTINIB | Targeted Therapy alone | 549 |
| BEVACIZUMAB | Targeted Therapy alone | 3 |
| BRIGATINIB | Targeted Therapy alone | 301 |
| CERITINIB | Targeted Therapy alone | 1229 |
| CRIZOTINIB | Targeted Therapy alone | 4023 |
| ERLOTINIB | Targeted Therapy alone | 6680 |
| GEFITINIB | Targeted Therapy alone | 7211 |
| LORLATINIB | Targeted Therapy alone | 325 |
| NINTEDANIB | Targeted Therapy alone | 1477 |
| OSIMERTINIB | Targeted Therapy alone | 455 |
| SORAFENIB | Targeted Therapy alone | 18 |
| SUNITINIB | Targeted Therapy alone | 3 |

Supplemental Table 2 – Regimen associated to their treatment intent

| **Chemotherapy regimen** | **Intent** |
|---|---|
| CARBOPLATIN | Palliative |
| CARBOPLATIN + DOCETAXEL | Palliative |
| CARBOPLATIN + ETOPOSIDE | Palliative |
| CARBOPLATIN + GEMCITABINE + PACLITAXEL | Palliative |
| CARBOPLATIN + IRINOTECAN | Palliative |
| CARBOPLATIN + NAB-PACLITAXEL | Palliative |
| CARBOPLATIN + PACLITAXEL | Palliative |
| CARBOPLATIN + PEMETREXED | Palliative |
| CARBOPLATIN + TOPOTECAN | Palliative |
| CARBOPLATIN + VINORELBINE | Palliative |
| CAV | Palliative |
| CISPLATIN | Palliative or Curative |
| CISPLATIN + DOCETAXEL | Palliative |
| CISPLATIN + DOXORUBICIN | Palliative |
| CISPLATIN + ETOPOSIDE | Curative |
| CISPLATIN + GEMCITABINE | Palliative |
| CISPLATIN + PACLITAXEL | Palliative |
| CISPLATIN + PEMETREXED | Palliative |
| CISPLATIN + VINORELBINE | Palliative |
| DOCETAXEL | Palliative |
| EC | Palliative or Curative |
| EP | Palliative or Curative |
| ETOPOSIDE | Palliative |
| EVEROLIMUS | Palliative |
| GEMCARBO | Palliative |
| GEMCITABINE | Palliative |
| PACLITAXEL | Palliative |
| PACLITAXEL + PEMETREXED | Palliative |
| PEMETREXED | Palliative |
| TOPOTECAN | Palliative |
| VINORELBINE | Palliative |

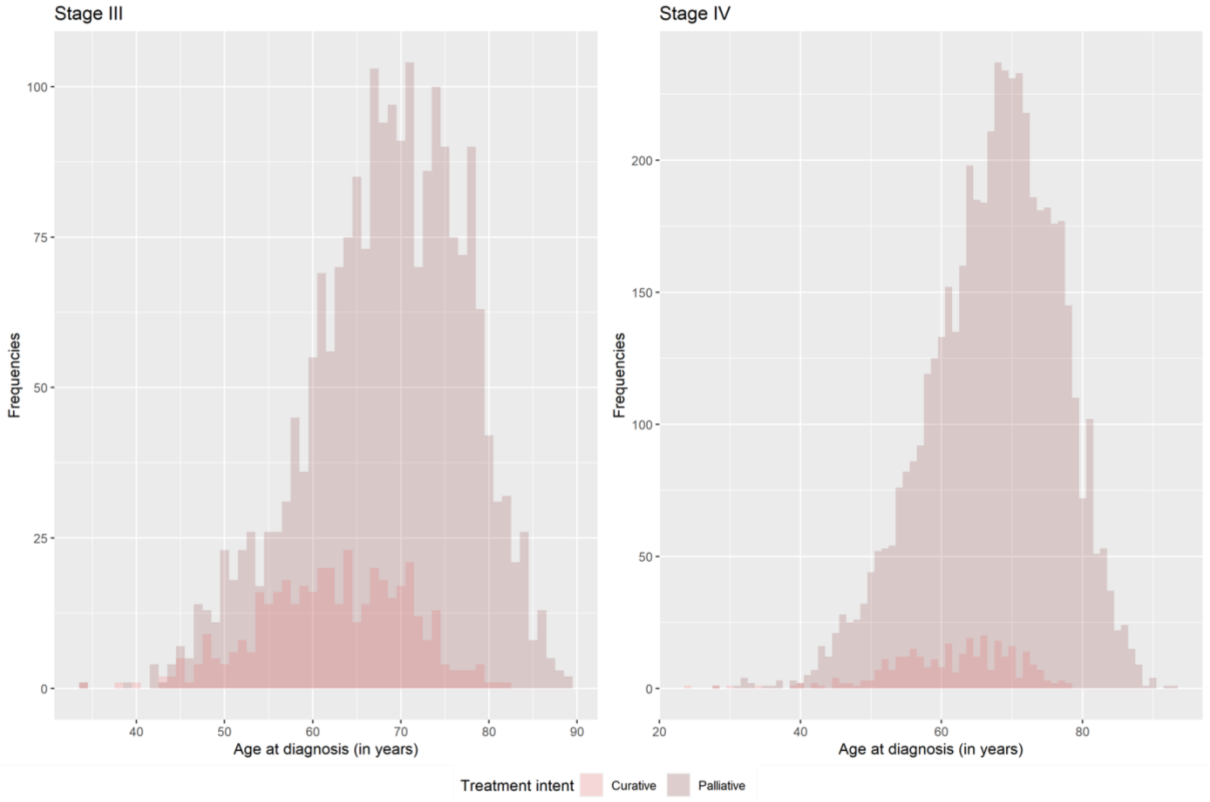


Supplemental Figure 1 – Age distribution by stage and treatment intent

Supplemental Table 3 - 30-and 90-day mortality rates and 6-and 12-month and median overall survival from the start of the 1st cycle of chemotherapy by stage and performance status in small cell lung cancer cases aged below and above 75 and treated with chemotherapy

| | Stage III | | Stage IV | |
|---|---|---|---|---|
| | **<75** | **≥75** | **<75** | **≥75** |
| **Performance status score = 0** | | | | |
| n | 465 | 91 | 633 | 123 |
| 30-day mortality - n (%) | 10 (2.2) | 2 (2.2) | 18 (2.8) | 6 (4.9) |
| 90-day mortality - n (%) | 19 (4.1) | 7 (7.7) | 69 (10.9) | 20 (16.3) |
| 6-month overall survival - % [95%CI] | 87.3 [84.3-90.4] | 80.2 [72.4-88.8] | 73.1 [69.8-76.7] | 58.5 [50.5-67.9] |
| 12-month overall survival - % [95%CI] | 66.4 [62.3-70.9] | 55.0 [45.6-66.2] | 36.5 [32.9-40.5] | 26.0 [19.3-35.1] |
| Median survival (months) [95%CI] | 16.5 [15.0-17.9] | 14.5 [10.6-18.2] | 9.3 [8.7-10.0] | 6.9 [6.1-8.4] |
| **Performance status score = 1** | | | | |
| n | 726 | 228 | 1436 | 371 |
| 30-day mortality - n (%) | 21 (2.9) | 5 (2.2) | 71 (4.9) | 21 (5.7) |
| 90-day mortality - n (%) | 45 (6.2) | 18 (7.9) | 192 (13.4) | 63 (17.0) |
| 6-month overall survival - % [95%CI] | 85.8 [83.3-88.4] | 80.6 [75.7-86.0] | 69.0 [66.7-71.4] | 62.8 [58.1-67.9] |
| 12-month overall survival - % [95%CI] | 58.7 [55.2-62.4] | 52.4 [46.3-59.3] | 29.9 [27.6-32.4] | 25.9 [21.8-30.7] |
| Median survival (months) [95%CI] | 14.4 [13.2-15.8] | 12.4 [10.8-14.5] | 8.3 [7.9-8.6] | 7.8 [6.9-8.3] |
| **Performance status score ≥2** | | | | |
| n | 355 | 168 | 1212 | 461 |
| 30-day mortality - n (%) | 24 (6.8) | 12 (7.1) | 173 (14.3) | 75 (16.3) |
| 90-day mortality - n (%) | 54 (15.2) | 31 (18.5) | 385 (31.8) | 184 (39.9) |
| 6-month overall survival - % [95%CI] | 72.4 [67.9-77.2] | 66.7 [60.0-74.2] | 45.6 [42.9-48.5] | 35.6 [31.5-40.2] |
| 12-month overall survival - % [95%CI] | 37.1 [32.4-42.5] | 32.1 [25.8-40.0] | 14.3 [12.4-16.4] | 10.2 [7.8-13.4] |
| Median survival (months) [95%CI] | 9.2 [8.4-10.4] | 8.1 [7.4-9.4] | 5.4 [5.1-5.9] | 4.3 [3.7-5.0] |
| **Missing performance status score** | | | | |
| n | 432 | 115 | 906 | 244 |
| 30-day mortality - n (%) | 20 (4.6) | 7 (6.1) | 80 (8.8) | 34 (13.9) |
| 90-day mortality - n (%) | 36 (8.3) | 14 (12.2) | 194 (21.4) | 68 (27.9) |
| 6-month overall survival - % [95%CI] | 80.6 [76.9-84.4] | 73.0 [65.4-81.6] | 59.8 [56.7-63.1] | 50.4 [44.5-57.1] |
| 12-month overall survival - % [95%CI] | 50.6 [46.1-55.5] | 41.3 [33.2-51.4] | 27.0 [24.2-30.0] | 18.4 [14.2-24.0] |
| Median survival (months) [95%CI] | 12.1 [11.3-13.4] | 9.7 [8.3-12.5] | 7.2 [6.8-7.6] | 6.1 [5.4-7.2] |

Note. CI: Confidence Interval
